# Epidemiological and clinical features of SARS-CoV-2 Infection in children during the outbreak of Omicron Variant in Shanghai, March 7-March 31, 2022

**DOI:** 10.1101/2022.04.28.22274421

**Authors:** Xiangshi Wang, Hailing Chang, He Tian, Jingjing Li, Zhongqiu Wei, Yixue Wang, Aimei Xia, Yanling Ge, Jiali Wang, Gongbao Liu, Jiehao Cai, Jianshe Wang, Qirong Zhu, Yanfeng Zhu, Xiaowen Zhai, Mei Zeng

## Abstract

**Objectives:** To understand the epidemiological and clinical characteristics of pediatric SARS-CoV-2 infection during the early stage of Omicron variant outbreak in Shanghai.

**Methods:** This study included local COVID-19 cases <18 years in Shanghai referred to the exclusively designated hospital by the end of March 2022 since emergence of Omicron epidemic. Clinical data, epidemiological exposure and COVID-19 vaccination status were collected. Relative risks (RR) were calculated to assess the effect of vaccination on symptomatic infection and febrile disease.

**Results:** A total of 376 pediatric cases of COVID-19 (median age:6.0±4.2 years) were referred to the designated hospital during the period of March 7-31, including 257 (68.4%) symptomatic cases and 119 (31.6%) asymptomatic cases. Of the 307 (81.6%) children;3 years eligible for COVID-19 vaccination, 110 (40.4%) received 2-dose vaccines and 16 (4.0%) received 1-dose vaccine. The median interval between 2-dose vaccination and infection was 3.5 (IQR: 3, 4.5) months (16 days-7 months). Two-dose COVID-19 vaccination reduced the risks of symptomatic infection and febrile disease by 35% (RR 0.65, 95% CI:0.53-0.79) and 33% (RR 0.64, 95% CI: 0.51-0.81). Two hundred and sixteen (83.4%) symptomatic cases had fever (mean duration: 1.7±1.0.8 days), 104 (40.2%) had cough, 16.4% had transient leukopenia; 307 (81.6%) had an epidemiological exposure in household (69.1%), school (21.8%) and residential area (8.8%).

**Conclusion:** The surge of pediatric COVID-19 cases and multiple transmission model reflect wide dissemination of Omicron variant in the community. Asymptomatic infection is common among Omicron-infected children. COVID-19 vaccination can offer protection against symptomatic infection and febrile disease.

## Introduction

The COVID-19 pandemic has caused devastation to the world’s population, resulting in more than 6 million deaths as of April 20, 2022. SARS-CoV-2 infection in most pediatric cases is mild as compared to adults and the direct effect on child health is limited [1]. However, the indirect impacts on child medical care, education and mental health are considerable owing to lockdown, disruption of essential health service delivery, prolonged school closure and isolation [2,3]. The continuous genetic evolution of SARS-CoV-2 virus results in the emergence of multiple new variants of concern (VOC), which are associated with enhanced transmissibility or increased virulence and immune escape [4]. The Omicron variant, which was detected in November 2021 and almost replaced Delta variant by the end of January 2022, has led to the fifth global wave of COVID-19 epidemic [5]. The significant rise of pediatric infection was reported in the United States with children aged <18 years, representing 17.0%-19.0% of all cases during the Omicron period since late December 2021 [6,7].

Pediatric COVID-19 cases only accounted for a small proportion of infection in the early stages of the COVID-19 pandemic when many countries implemented non-pharmaceutical interventions and strict containment measures [8-12]. However, the incidence rate of COVID-19 in children showed a rising trend in the epidemic countries following suspension of lockdown and school reopening [12,13]. After the large-scale epidemic in early 2020, China entered a normalization stage of prevention and control, and massive COVID-19 vaccination campaign was launched nationwide in 2021. Inactivated SARS-CoV-2 vaccine BBIBP-CorV (by Sinopharm) and CoronaVac (by Sinovac) were approved for emergency use in children 3-17 years on June 2021 and COVID-19 vaccination program was initiated in pediatric population since late July 2021 across China. From May 2020, local pediatric COVID-19 infection linked to sporadic and cluster transmission were occasionally reported in China until the community outbreak of Omicron variant appeared in Hong Kong Special Administrative Region since January 6, 2022 and subsequently in Shanghai since early March 2020 [14]. Omicron variant spread rapidly in Shanghai by the end of March and led to a surge of pediatric COVID-19 cases citywide. Here, we describe epidemiological and clinical characteristic of Omicron variant infections in Shanghainese children during the early stage of the outbreak.

## Subject and Method

### Subject

In this study, we included local COVID-19 cases <18 years of age who were notified in Shanghai and admitted to the exclusively designated hospital in Shanghai by the end of March 2022. Prior to 28 March, all pediatric COVID-19 cases notified in Shanghai were referred to the designated hospital for concentrating management and isolation. Since 28 March when a large number of cases were confirmed by massive screening test, most of asymptomatic and mild pediatric cases aged 5-17 years were almost sent to Fangcang shelter hospitals. All confirmed cases irrespective of symptoms are required for in-hospital isolation and are discharged if the cycle threshold (Ct) value for the viral nucleic acid is great than 35 on PCR test for the two consecutive respiratory samples taken 24-hour apart [15].

### Case definition and classification

All COVID19 cases were laboratory-confirmed by the Shanghai CDC reference laboratory using real-time RT-PCR commercial kit. The Ct value <40 was defined as a positive nucleic acid amplification test. COVID-19 cases were classified as asymptomatic and symptomatic cases. Symptomatic cases were classified as mild, moderate and severe cases. An asymptomatic case is defined as a person with a positive nucleic acid test but without any clinical symptom of COVID-19. A confirmed symptomatic case is defined as a person presenting clinical signs and symptoms of COVID-19. COVID-19 disease severity classification is based on the WHO guidance [16]. Pneumonia was diagnosed based on clinical signs (fever and/or cough accompanying with one of the following signs: moist rales on auscultation, difficulty breathing/dyspnoea, fast breathing, chest indrawing), radiological findings compatible with pneumonia.

### Data collection

Data were collected via a face-to-face interview with parents or teenagers and electronic medical chart, including: demographic information, epidemiological exposure setting, COVID-19 vaccination status on dose and date, clinical symptoms, laboratory findings and chest imaging if examined, treatment and outcome. Informed consent from parents was not required by the ethics committee because all data were de-identified and not involved in personal privacy.

### Statistical analysis

Data was entered into Excel version 2016 (Microsoft, Redmond, Washington) for analysis and the statistical analysis was preformed using SPSS (IBM Statistic 23.0). Categorical variables are described as absolute numbers and percentage. Continuous variables with normal distribution are expressed as mean ± standard deviation. Median (25% to 75% interquartile range (IQR) are used when the frequency distributions were skewed. Differences between groups are compared using Mann-Whitney *U*-test and Student’s *t*-test as appropriate. A difference with P <0.05 is considered to be statistically significant. Relative risks (RR) were calculated using proportion of symptomatic infection and febrile cases by vaccination status, with the referent group being ≥2-dose vaccinees.

## Results

### Demographic characteristics

The first local pediatric case was notified on March 2022 and increased remarkably from 14 March onwards (as shown in Figure 1). As of 31 March, a total of 376 pediatric cases of COVID-19 were referred to the exclusively designated hospital. The ratio of male-to-female was 1.1 (206/170). The 376 cases were aged 11 days-17 years with the median age of 5.0 years (IQR: 2, 9) and the mean age of 6.0±4.2 years: 28 (7.4%) cases in age group <1 year, 76 (20.2%) cases in age group 1-2 years, 94 (25.0%) cases in age group 3-5 years, 134 (35.6%) cases in age group 6-11 years, and 44 (11.7%) cases in age group ≥12 years.

**Figure 1.**
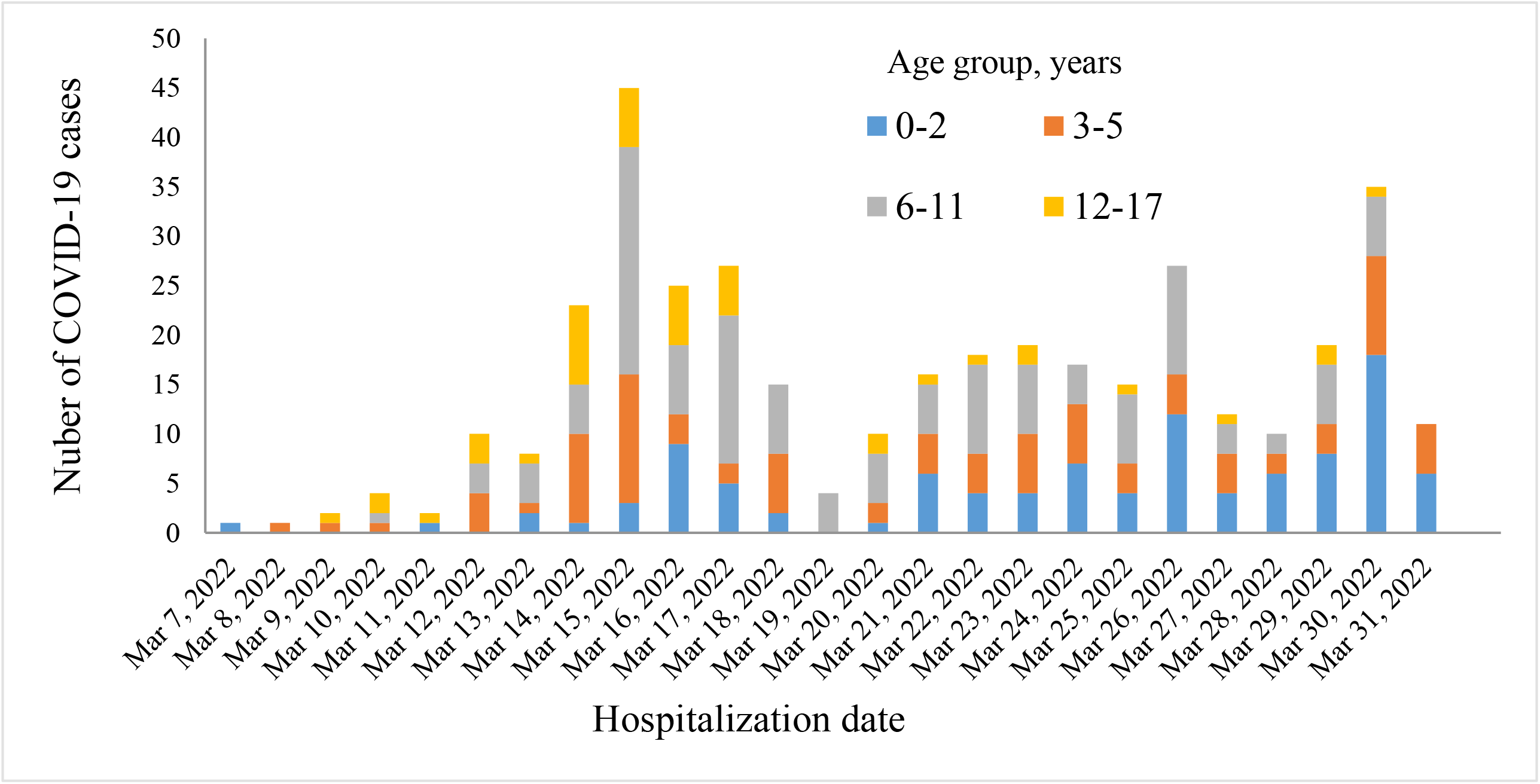
Daily COVID-19 cases referred to the designated hospital for children aged <18 years from March 1 to March 31, 2022

### Epidemiological exposure

Three hundred and seven (81.6%) cases had a clear history of exposure, of whom, 213 (69.1%) had a close contact with confirmed adult cases in household, 67 (21.8%) had a clear contact with confirmed child cases in school, and 27 (8.8%) had an epidemiological linkage to residential area where cluster cases of COVID-19 were reported. As shown in Figure 2, the first child case acquired infection in family, soon after, child cases linked to possible community transmission were found, who had no clear exposure.

**Figure 2.**
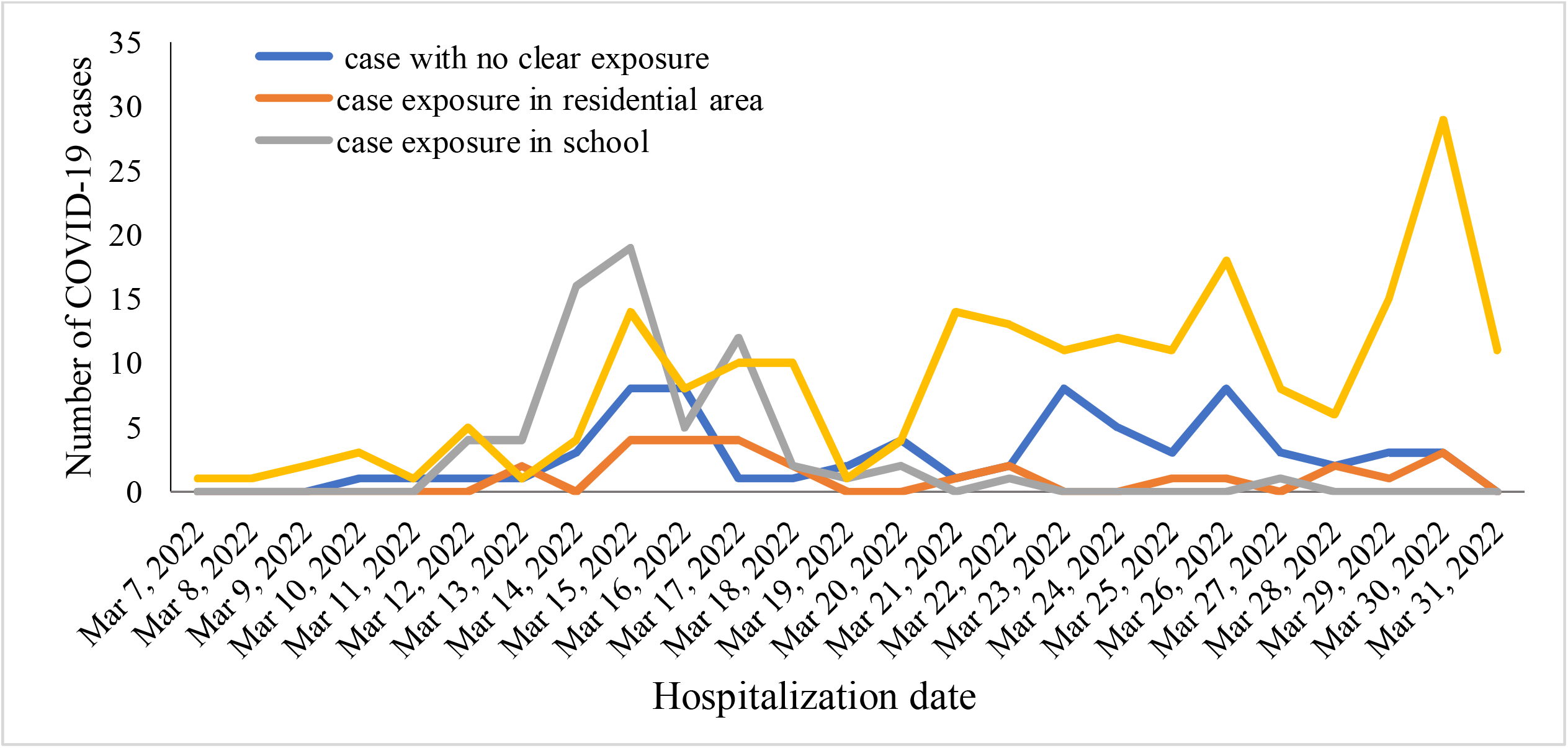
Model of epidemiological exposure over time among pediatric COVID-19 cases

### Vaccination status

A total of 126 had received at least one dose of an inactivated COVID-19 vaccine, accounting for 33.5% of the total 376 pediatric cases and 46.3% of the 272 pediatric cases aged ≥3 years eligible for COVID-19 vaccination in China. Of the 272 vaccine-eligible children, 146 (53.6%) were unvaccinated, 110 (40.4%) had received 2 doses and 16 (4.0%) had received 1 dose. Among the 94 preschool children aged 3-5 years, the proportions of 1 dose and 2 doses of COVID-19 vaccination were 3.2% (3/94) and 5.3% (5/94), respectively. Among the 178 school children aged 6-17 years, the proportions of 1 dose and 2 doses of COVID-19 vaccination were 7.3% (13/174) and 59.0% (105/174), respectively. Overall, the interval between vaccination and breakthrough infection ranged from 16 days to 7 months (median: 3.5 (IQR: 3, 4.5) months).

As shown in table 1, 2-dose COVID-19 vaccination reduced the risk of symptomatic infection and febrile disease by 35% (0.65, 95% CI: 0.53-0.79) and by 36% (RR 0.64, 95% CI: 0.51-0.81) in children 0-17 years, by 29% (RR 0.71, 95% CI: 0.57-0.88) and 29% (RR 0.71, 95% CI: 0.55-0.92) in children 3-17 years eligible for COVID-19 vaccine. However, one-dose vaccination could not significantly decrease the relative risks of symptomatic infection and febrile disease.

**Table 1.**
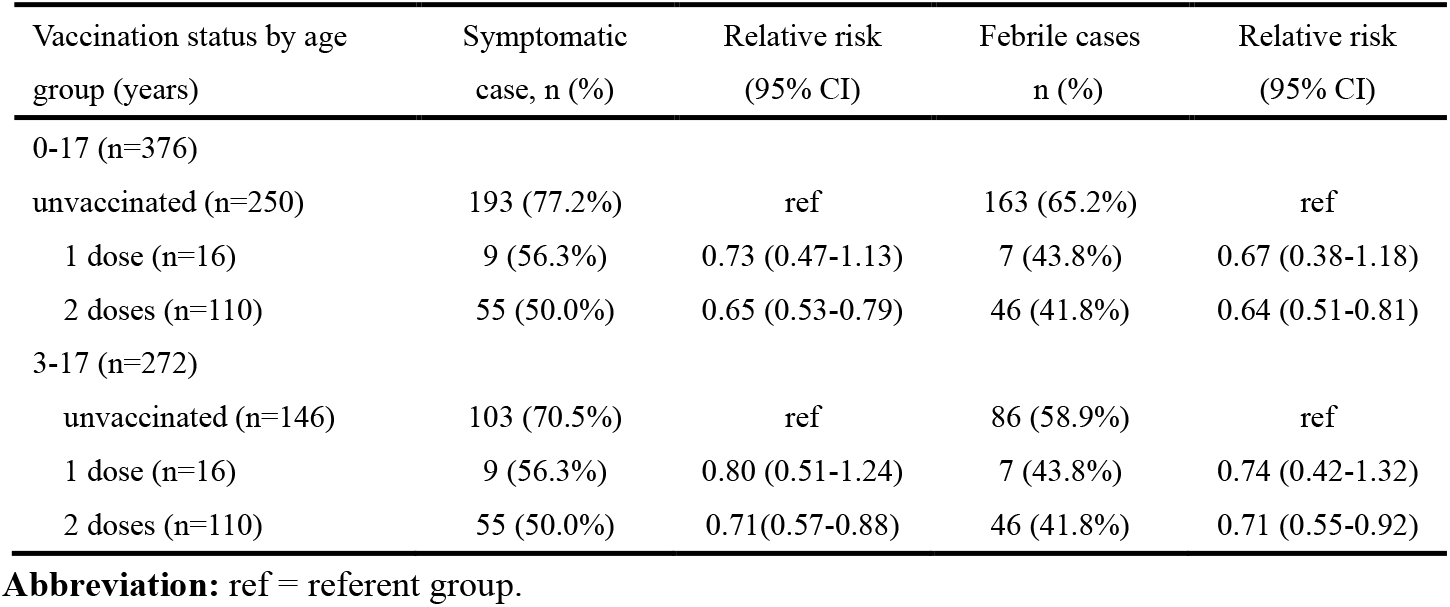
Clinical characteristics of SARS-CoV-2 infection according to COVID-19 vaccination status

**Table 2.** Clinical characteristics of SARS-CoV-2 virus infection by age group

| Clinical characteristics | Total<br>(n=376) | Age group (years) |  |  | *P* value | Vaccination status |  | *P* value |
| --- | --- | --- | --- | --- | --- | --- | --- | --- |
|  |  | <3 | 3-5 | 6-17 |  | Unvaccinated | 2-dose vaccination |  |
|  |  | (n=104) | (n=94) | (n=178) |  | (n=250) | (n=110) |  |
| Asymptomatic cases, n (%) | 119<br>(31.6%) | 14<br>(13.5%) | 29<br>(30.9%) | 76<br>(42.7%) | 0 | 57 (22.8%) | 55 (50.0%) | 0 |
| Symptomatic cases, n (%) | 257<br>(68.4%) | 90<br>(86.5%) | 65<br>(69.1%) | 102<br>(57.3%) | 0 | 193 (77.2%) | 55 (50.0%) | 0 |
| Febrile cases, n (%) | 216<br>(57.4%) | 44<br>(42.3%) | 59<br>(62.8%) | 80<br>(44.9%) | 0.006 | 163 (65.2%) | 46 (41.8%) | 0 |
| Fever spike (°C), mean±SD | 38.9±0.6 | 39.0±0.7 | 39.0±0.6 | 38.8±0.6 | 0.064 | 38.9±0.6 | 38.8±0.7 | 0.27 |
| Fever duration (days), mean±SD | 1.7±0.8 | 1.7±0.9 | 1.6±0.6 | 1.8±0.8 | 0.45 | 1.6±0.8 | 1.9±0.9 | 0.19 |

### Clinical manifestation and course

Of the 376 cases, 257 (68.4%) presented symptoms and 119 (31.6%) had no symptoms before and duration hospitalization. Of the 257 symptomatic cases, 216 (83.4%) experienced fever (axillary temperature >37.5°C) with a mean fever spike of 38.9± 0.6°C (range: 37.6-41°C) and a mean fever duration of 1.7±1.0.8 days (range: 0.5-4 days), 104 (40.2%) presented cough, 28 (10.8%) self-reported sore throat, 13 (5.0%) self-reported stuffy nose, 6 (2.3%) had runny nose, 11 (4.2%) had nausea or vomiting or diarrhea, 2 (0.8%) self-reported transient loss of taste and smell. No severe case was diagnosed. Twenty five cases had chest CT performed due to fever >38.5°C lasting for 3 days or cough worsening after admission or routine examination prior to the referral. The chest images showed patchy infiltrates or ground-glass opacity in 4 cases and one of them was right lung lobar pneumonia caused by Mycoplasma pneumoniae. Six (1.6%) cases had comorbidity including brain tumor, febrile seizure, psychomotor retardation, hemophilia, Henoch-Schonlein purpura, and cardiac arrhythmia in each.

As shown in table 2, 22.8% (57/250) of unvaccinated cases were asymptomatic while 50.0% (55/110) of 2-dose vaccinated cases were asymptomatic (P=0.000); 65.2% (163/250) of unvaccinated cases were febrile while 41.8%% (46/110) of 2-dose vaccinated cases were febrile (P=0.000). Symptomatic infection was significantly frequently seen in the age group <3 years than in the age group 3-5 years (P=0.003) and 6-17 years (P=0.000). Fever was significantly frequently seen in the age group 3-5 years than in the age group <3 years (P=0.000) and 6-17 years (P=0.005).

Of the 225 case who had complete peripheral blood cell count tested, 37 (16.4%) had white blood cell (WBC) count <4×10^9^/L, 173 (76.9%) had WBC count 4-9×10^9^/L, 13 (5.8%) had WBC count 10-14×10^9^/L and 2 (7.1%) had WBC count ≥15×10^9^/L. The WBC count ranged from 1.9×10^9^/L to 15.5×10^9^/L. No thrombopenia was observed. Of the 187 cases who had peripheral blood C-reactive protein (CRP) tested, 178 (95.2%) had CRP <8 mg/L, 8 (4.3%) had CRP >8 mg/L (range: 8.8-35.8 mg/L) and 1 (0.5%) had CRP 56 mg/L who had co-infection with *mycoplasma pneumoniae* and developed typical lobar pneumonia in right lung. Of the 196 cases who had serum biochemical markers and 8 (4.1%) showed slightly elevated liver enzyme.

For symptomatic cases, Ibuprofen and or Chinese traditional medicines were prescribed depending on the personalized condition and medication compliance. Only one case who had a clear diagnosis of mycoplasma pneumonia was prescribed antibiotics. All cases were discharged when the Ct value of the nucleic acid of SARS-CoV-2 virus reached >35. The average duration of Ct value of the nucleic acid of SARS-CoV-2 virus >35 since admission was 11.7±3.7days (range: 3-25 days; symptomatic verse asymptomatic: 11.7±3.6 verse 11.7±3.9, P=0.064).

## Discussion

This study first presents the epidemiological and clinical profiles of Omicron variant infection in localized children during the early phase of outbreak in Shanghai. As of 31 March 2022, all pediatric COVID-19 cases were mild (68.4%) or asymptomatic (31.6%). However, a few of severe pediatric cases were reported during the period of COVID-19 outbreak in Wuhan in early 2020 [17]. Moreover, the proportion of asymptomatic cases was 2-time more than that seen in the Wuhan outbreak. We reason that high coverage of COVID-19 vaccination among Shanghainese children is very likely to lower the risk of severe Omicron infection-associated disease. The mass COVID-19 vaccination roll-out among children 3-17 years started between Mid-Aug 2021 and December 2021 in Shanghai and the estimated coverage rate of 2-dose COVID-19 vaccination among children 3-17 years has exceeded 70% by of the end of March in 2022. In this case cohort 46.3% of children eligible for COVID-19 vaccination prior to 16 days to 7 months (median: 3.5months). Observational studies from some countries with the high levels of population immunity generated by natural infection or vaccine have shown receipt of two doses of COVID-19 vaccines and a booster dose can offer protection against symptomatic and severe Omicron infection in a short-term period of vaccination [4,18-22].

Current evidences consistently show a reduction in neutralizing antibody against Omicron in serum of convalescent or vaccinated individuals, resulting in Omicron’s immune escape potential against vaccine- and infection-induced immunity [4,23]. However, two recent study based on real-world observation among children showed the modest effectiveness for COVID-19 vaccine against Omicron infection [25,26]. Based on our findings, receipt of 2-dose inactivated COVID-19 vaccine within 17 days to 7 months after fully primary vaccination potentially reduced the risk of symptomatic Omicron infection by 31% and febrile disease by 59% in children. We did not estimate vaccine protection against severe infection because no severe COVID-19 cases were diagnosed. There is also evidence of waning of vaccine effectiveness over time of the primary series against infection and symptomatic disease for the studied vaccines. However, the vaccine effectiveness against Omicron infection and disease can be restored and increase to > 40% to 80% within a short follow-up time after a third booster dose in studies from five countries (United Kingdom, Denmark, Canada, South Africa, USA) [4]. Ten cases of reinfection with Omicron variant were identified within 23 to 87 days of a previous Delta infection was reported in the USA and most were pediatric cases [26]. Thus, eligible children and adolescents should remain up to date with recommended COVID-19 vaccination in response to Omicron outbreak. So far, a third booster dose of COVID-19 vaccine has been recommended for use in adults but not in children in China. In light of the field findings, a booster dose should also be recommended for eligible children 3-17 years.

We observed that school children aged 6-11 years comprised the most cases, followed by home-care children <2 years and preschool children 3-5 year. The distribution of age groups in the early stage of outbreak reflects the cluster transmission of COVID-19 centered in elementary school, kindergarten, and household. Of note, age group 12-17 years accounted for the smallest proportion of pediatric cases, among which, the high coverage rate of COVID-19 vaccination was as high as 95%. Based on the epidemiological investigation, more than 80% children had a clear history of exposure, mostly occurring in family (69.1%) and school (21.8%), occasionally in residential area (8.8%). The remaining 18.4% of children had no clear contact with confirmed cases, reflecting small-scale community transmission had already appeared prior to the large-scale outbreak since April. During the 2020 outbreak of COVID-19, 80%-90% of confirmed child cases were family cluster cases and community transmission was unusual in China [17,27]. Rapid increases in pediatric COVID-19 cases and epidemiological unrelated cases also suggest the occurrence of high community transmission of Omicron variant in Shanghai since the early epidemic wave.

We observed most of localized pediatric cases (83.4%) of symptomatic Omicron infection presenting fever. However, fever is less commonly seen in pediatric COVID-19 cases reported in China (58%) and the USA (56%) during the first wave of pandemic in 2020 [10, 17]. Fever could be helpful for early recognition and diagnosis of COVID-19 because parents always worry about the febrile child and visit hospital for seeking medical care. The febrile course of Omicron infection is brief with a mean fever duration of 1.7±1.0.8 days, significantly shorter than fever duration seen influenza (4 days) [28]. The febrile duration is helpful to differentiate COVID-19 from influenza in children when the epidemics of COVID-19 and influenza overlap.

The potential role in transmission for most asymptomatic and mild child cases should not be neglected. A study showed that symptomatic and asymptomatic children can carry high quantities of live SARS-CoV-2, creating a potential reservoir for transmission [29]. Vaccinees with mild or asymptomatic Omicron infection shed infectious virus 6-9 days after onset or diagnosis, even after symptom resolution [30]. In fact, asymptomatic infection in children was underestimated in the early stage of outbreak because massive screening of COVID-19 cases had not been carried out before 28 March. After citywide large-scale screening, notifiable asymptomatic cases accounted for 90% more or less in April. Asymptomatic infection was much more common in vaccinated children than in unvaccinated children (50% vs 22.8%). Vaccination can offer protection against symptomatic infection and febrile disease, on the other hand, the role of asymptomatic children play in viral transmission is of attention during outbreak. High prevalence of asymptomatic infection is likely a major factor in the widespread of the Omicron variant among population.

In summary, COVID-19 is mild and subtle in Shanghainese children with the high level of vaccine-induced immunity during the early stage of Omicron outbreak. COVID-19 vaccination can offer partial protection against symptomatic COVID-19. Ongoing Omicron epidemic will increase the risk of exposure among children with underlying medical conditions, who are usually unvaccinated, therefore, severe COVID-19 infection is anticipated to be encountered in children. Non-pharmaceutical interventions in combination with vaccination strategies are critical to prevent infection and severe disease and to mitigate the impact of COVID-19 in pediatric population.

### Transparency declaration

The authors have declared that there are no conflicts of interest in relation to this work. Disclosure forms provided by the authors are available as Supplementary material.

## Data Availability

The datasets analyzed during the present study are available from the corresponding author upon reasonable request

## Funding

This work was supported by the Science and Technology Commission of Shanghai Municipality (NO. 20JC141020002), the Key Development Program of Children’s Hospital of Fudan University (EK2022ZX05), the Three-Year Action Plan for Strengthening the Construction of Public Health System in Shanghai (2020-2022) (GWV-3.2), and the Key Discipline Construction Plan from Shanghai Municipal Health Commission (GWV-10.1-XK01). None of the funding organizations played any role in the trial design, data collection, analysis of results, or writing of the manuscript.

## Author contributions

Conceptualization: YZ, XZ, MZ; Methodology: XW, HC, HT, MZ; Software: XW, HC, HT; Validation: JC; Formal Analysis: XW, HC, HT, YZ, MZ; Investigation: JL, ZW, YW, AX, YG, JW, GL, JC; Writing-Original Draft: XW, HC, YZ, MZ; Writing-Review & Editing: XW, MZ; Visualization: JC, HC; Supervision: YZ, GL, JW, QZ, XZ, MZ; Project Administration: XZ; Funding acquisition: XZ, MZ. All authors approved the manuscript for publication.

## Notes

### Competing Interest Statement

The authors have declared no competing interest.

### Funding Statement

This study was funded by the Science and Technology Commission of Shanghai Municipality (NO. 20JC141020002), the Key Development Program of Children's Hospital of Fudan University (EK2022ZX05), the Three-Year Action Plan for Strengthening the Construction of Public Health System in Shanghai (2020-2022) (GWV-3.2), and the Key Discipline Construction Plan from Shanghai Municipal Health Commission (GWV-10.1-XK01). None of the funding organizations played any role in the trial design, data collection, analysis of results, or writing of the manuscript.

### Author Declarations

Ethics committee/IRB of Children's Hospital of Fudan University waived ethical approval for this work

